# A clinical observational analysis of aerosol emissions from dental procedures

**DOI:** 10.1101/2021.06.09.21258479

**Authors:** T. Dudding, S. Sheikh, F. Gregson, J. Haworth, S. Haworth, B.G. Main, A.J. Shrimpton, F.W. Hamilton, AERATOR group, A.J. Ireland, N.A. Maskell, J.P. Reid, B.R. Bzdek, M. Gormley

## Abstract

Aerosol generating procedures (AGPs) are defined as any procedure releasing airborne particles <5 μm in size from the respiratory tract. There remains uncertainty about which dental procedures constitute AGPs. We quantified the aerosol number concentration generated during a range of periodontal, oral surgery and orthodontic procedures using an aerodynamic particle sizer, which measures aerosol number concentrations and size distribution across the 0.5–20 μm diameter size range. Measurements were conducted in an environment with a sufficiently low background to detect a patient’s cough, enabling confident identification of aerosol. Phantom head control experiments for each procedure were performed under the same conditions as a comparison. Where aerosol was detected during a patient procedure, we assessed whether the size distribution could be explained by the non-salivary contaminated instrument source in the respective phantom head control procedure using a two-sided unpaired t-test (comparing the mode widths (log(*σ*)) and peak positions (D_P,C_)). The aerosol size distribution provided a robust fingerprint of aerosol emission from a source. 41 patients underwent fifteen different dental procedures. For nine procedures, no aerosol was detected above background. Where aerosol was detected, the percentage of procedure time that aerosol was observed above background ranged from 12.7% for ultrasonic scaling, to 42.9% for 3-in-1 air + water syringe. For ultrasonic scaling, 3-in-1 syringe use and surgical drilling, the aerosol size distribution matched the non-salivary contaminated instrument source, with no unexplained aerosol. High and slow speed drilling produced aerosol from patient procedures with different size distributions to those measured from the phantom head controls (mode widths log(σ)) and peaks (D_P,C_), *p*< 0.002) and, therefore, may pose a greater risk of salivary contamination. This study provides evidence for sources of aerosol generation during common dental procedures, enabling more informed evaluation of risk and appropriate mitigation strategies.

## Introduction

Transmission of respiratory diseases, such as severe acute respiratory syndrome coronavirus 2 (SARS-CoV-2), the causative virus for the coronavirus disease 2019 (COVID-19), can occur through direct or indirect physical contact, droplet inhalation or airborne transmission [1]. Aerosols and droplets are created when the surface tension of a fluid is overcome by force, for example from air turbines within dental drills [2]. High viral loads present in the course of COVID-19 infection make dental aerosols a plausible source of infective particles [2-4]. A recent study has demonstrated that asymptomatic patients attending dental care settings can be positive for SARS-CoV-2 [5]. Understanding and managing the disease risk posed by dental aerosols is important to protect patients and dental teams.

Aerosol generating procedures (AGPs) may result in respiratory disease transmission, and are defined as any procedure that can result in the release of airborne particles <5 μm in size from the respiratory tract of an individual [6]. Recent work has shown that dental AGPs generate particles with a size distribution sufficiently wide to potentially incorporate SARS-CoV-2 virions (0.05–0.15 μm) [7]. Aerosol generated during dental procedures is of respirable size, can remain suspended in air around the dental team [8], and is removed primarily by ventilation parameters of the room. By contrast, larger splatter particles (>50 μm), often follow a ballistic trajectory, and are rarely detected more than 2 m from the source during dental procedures [9].

Potential AGPs have attracted additional mandatory infection control practices, including personal protective equipment (PPE), ensuring adequate ventilation and allowing additional ‘fallow’ time between patients to enhance aerosol dispersion [10]. In dentistry there remains uncertainty about which procedures constitute an AGP, with a recent systematic review highlighting this evidence gap [11]. Previous studies suffer from inherent drawbacks, using instruments with limited sensitivity to accurately detect aerosol, such as culture or settling plates, which do not account for the suspension of particles or those removed through ventilation [12-14]. Others have employed simulation on phantom heads [15, 16], which may not accurately capture the real clinical scenario. Some recent studies have used high-resolution electrical low-pressure impactor particle sizers [7] and portable scanning mobility particle sizers [17], to capture the smaller aerosol particles (<10 μm) likely produced during dental AGPs. However, this work has also been performed in phantom heads rather than on dental patients.

For any dental procedure on a patient, there are three aerosol sources to consider. First, the host (patient) aerosol generated during breathing, speaking, or coughing may be infectious to the dental team in close proximity [18]. The second source of aerosol is the instrument generated aerosol, which is not considered infectious as there is no physical interaction with the host. Finally, there is salivary-contaminated aerosol generated by the action of the instrument in a potentially infectious host, which might be infectious. One challenge is separating this salivary-contaminated aerosol from the non-salivary contaminated instrument source. In this study we quantified the aerosol number concentration, in the 0.5–20 μm size range, produced during a wide range of dental procedures in a real-world clinical setting. We also aimed to determine whether aerosol detected was intrinsically generated from the non-salivary contaminated dental instrument, or was likely to be contaminated using aerosol size distribution analysis and modelling with phantom head controls. These measurements were conducted in an environment with an aerosol background concentration low enough to reliably detect a patient’s cough, allowing robust detection of any aerosol generated during dental procedures [19].

## Methods

### Ethical approval and patient recruitment

This study obtained ethical approval as part of the AERosolisation And Transmission Of SARS-CoV-2 in Healthcare Settings (AERATOR) study via the Northwest Research Ethics Committee (Ref: 20/NW/0393) and was conducted in accordance with the STROBE guidelines (Strengthening the Reporting of Observational Studies in Epidemiology). Adult patients >18 years old on waiting lists requiring either periodontal, oral surgery or orthodontic treatment were recruited consecutively. Each patient was contacted via telephone, received an information leaflet via post, and provided written consent on the day of treatment.

### Environment and equipment

An Aerodynamic Particle Sizer (APS) (TSI Incorporated, model 3321, Shoreview, NM, USA; detection range: 0.5-20 μm diameter particles) was used to measure aerosol. A custom 3D-printed funnel (RAISE3D Pro2 Printer, 3DGBIRE, Chorley, UK) made from polylactide, with a maximum diameter of 150 mm, cone height of 90 mm and a 10 mm exit port, was attached to the APS inlet using conductive silicone sampling tubing (TSI, 3001788), approximately 0.90 m long and 4.80 mm in diameter. The experimental set-up is shown in **Supplementary Fig. 1**. The APS was set to sample aerosol number concentration once per second. Further detail on the environment and instruments used can be found in the **Supplemental Material**.

### Baseline patient measurements

Baseline readings were taken from each participant including tidal breathing at rest (60 s), counting out loud (60 s) and three voluntary coughs. The funnel was positioned at source (as close to the mouth as possible), with the patient seated upright [20]. Baseline characteristics of patients were reported using median and range for continuous data, alongside counts and percentages for categorical data, stratified by specialty type. To assess differences among specialties, age and sex distribution was compared using one-way ANOVA and Fisher’s exact test respectively.

### Patient dental procedure aerosol measurement

We conducted an initial pilot study to investigate the optimum position and orientation for the 3D-printed funnel when sampling dental aerosol. This was determined to be 22 cm from soft tissue nasion to the top of the funnel, at approximately 45 degrees on the patient’s left side (11 o’clock position). For every case, a full mouth examination was carried out using a dental mirror, followed by local anaesthetic administration when indicated. Each patient received 3-in-1 syringe air drying (30 s), water (30 s) and then combined air and water (30 s) applied to their all their teeth. When necessary, up to 3-minute intervals between procedural steps were allowed for background reading levels to stabilise. The remainder of the treatment session was dictated by clinical need. A detailed description of the treatments and time-stamped protocols are provided in the **Supplementary Methods**.

### Phantom head control procedure aerosol measurement

To measure aerosol generated by the dental instruments alone, we conducted high fidelity control experiments in triplicate, in a phantom head unit. For phantom head control data, the aerosol number concentration and size distribution were extracted for further analysis. Further detail can be found in the **Supplementary Methods**.

### Statistical analysis

#### Total procedure aerosol number concentration

The aerosol number concentration for each procedure and baseline measurement were compared by calculating particle number concentration detected above background for each patient (irrespective of particle size). As the length of procedure differed across patients, we sampled the mean particle number concentration across the sampling time for each patient and the per patient values were combined to give median and inter-quartile ranges of total aerosol number concentration for each procedure.

#### Procedure aerosol size distributions

The aerosol size distributions from the phantom head control and patients were compared, with the assumption that if the distributions were the same, all aerosol detected from the patient during the procedure could be explained by the non-salivary contaminated instrument source (represented by the phantom control). For each procedure, mean aerosol number concentrations (dN) for a range of particle size bins (D_p_) were calculated by averaging across patients. These were transformed (dN/dLog(D_p_)/cm^-3^) in order to normalise the data, enabling visual comparison of the size distribution in a standardised form typical for reporting aerosol size distributions (**Fig. 1**). For each procedure, the shape of the phantom head control and patient size distributions were compared visually.

**Figure 1.**
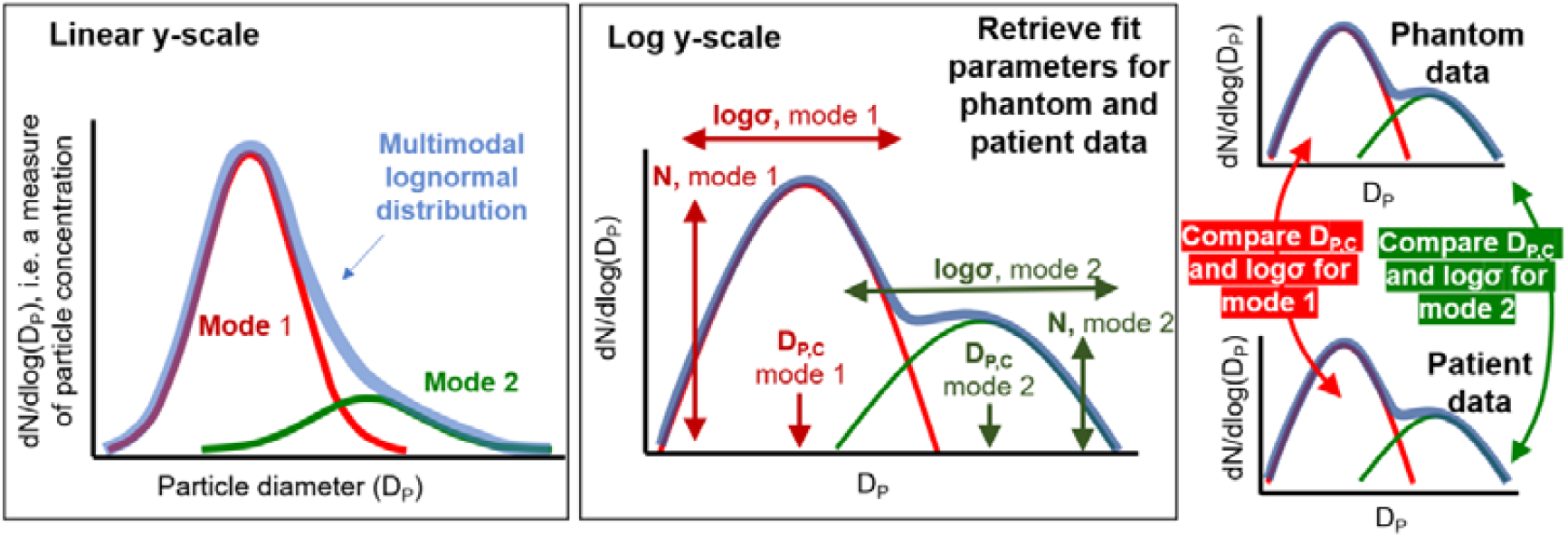
Description of the parameters used to describe the average size distribution detected above background. The mode width is described by log sigma and the peak of the mode by D_P,C_. Mode amplitude parameter (N) was not compared, as it is highly dependent on sampling variability.

In the phantom head control, the model best describing the size distribution (highest r^2^) was identified by iteratively altering the number of modes (uni-modal, bi-modal, or tri-modal) using **Supplementary Equation S1**. Once fitted, mode parameters (N, D_P,C_, log(*σ*)) were compared between patient and phantom head control as illustrated in **Fig. 1**. Aerosol size distributions inherently provide a robust fingerprint of source with different aerosol sources yielding different log-normal distributions with different mean sizes and breadths/standard deviations. This enabled us to attribute aerosols to specific sources e.g., phantom head or patient [21].

To further assess if size distributions between the phantom head control and patient differed other than by chance, a two-sided unpaired t-test was used to compare the mode widths (log(*σ*)) and peak positions (D_P,C_). The mode amplitude parameter (N) was not compared as it is highly dependent on variabilities in sampling efficiency, for example direction of the exhaled airflow, which is not comparable across phantom head and patient sampling. After accounting for multiple parameters compared across instruments (n= 26), a Bonferroni adjusted *p*-value of 0.002 was used.

## Results

Forty-one patients were included in the study with a median age of 47 years (range: 18–75) (**Supplementary Tables 1 & 2**). The mean background aerosol number concentration across patients was 0.18 (+/-SD 0.12) particles cm^-3^ (180 particles per litre). In total, fifteen dental procedures were assessed during periodontal, orthodontic, and oral surgery treatments. Of these, examination with a dental probe, hand scaling, local anaesthetic delivery, routine extraction (with forceps and/or elevator), raising a soft tissue flap, orthodontic bracket removal, alginate impression taking, 3-in-1 water only, and suturing did not produce any aerosol (**Supplementary Table 3**). For the other six procedures where aerosol was detected, the percentage of total procedure time that aerosol was observed was 12.7% for ultrasonic scaling, 24.8% for 3-in-1 air only, 75.3% for 3-in-1 air + water, 40.1% for high-speed drilling, 49.9% for slow speed drilling and 55.6% for surgical drilling (**Table 1**).

**Table 1.** Dental procedures for which aerosol was detected above background.

| Procedure | Number of patients* | Total sampling time for procedure (s) | Time aerosol detected above background (s) | Percentage time aerosol detected above background (%) |
| --- | --- | --- | --- | --- |
| Ultrasonic scaling | 12 | 12,272 | 1,559 | 12.7 |
| 3-in-1 air only | 35 | 801 | 199 | 24.8 |
| 3-in-1 air + water | 33 | 772 | 581 | 75.3 |
| High speed drilling | 15 | 3,849 | 1,543 | 40.1 |
| Slow speed drilling | 15 | 3,324 | 1,632 | 49.9 |
| Surgical drilling | 9 | 568 | 316 | 55.6 |

\* Some procedures were conducted in more than one patient

### Aerosol number concentrations from dental procedures

The aerosol number concentration for each procedure is shown in **Fig. 2**. Participant breathing and speaking had similar number concentrations and size distributions to background aerosol, indicating the background dominated the signal for these activities. High speed drilling produced 10-fold more aerosol (median 118.38 cm^-3^) compared to the other five procedures (ultrasonic scaling, surgical drilling, 3-in-1 syringe air/ air + water, and slow speed drilling), which were comparable with median number concentrations of approximately 10 cm^-3^.

**Figure 2.**
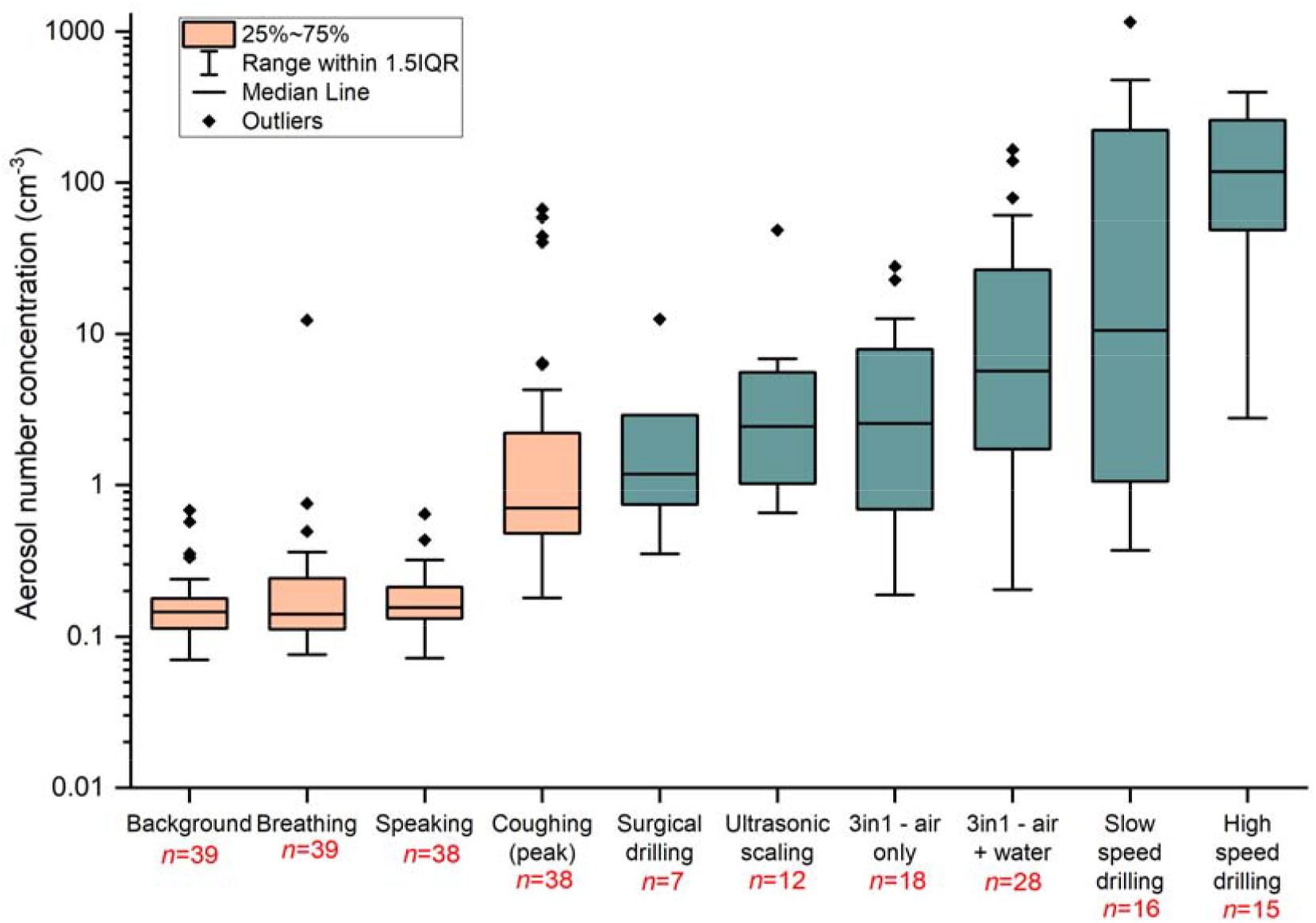
Box and whiskers plot of total aerosol number concentration for baseline measurements (orange) and dental procedures for which aerosol was detected above mean background value (green). The aerosol number concentration is reported on a logarithmic scale.

### Aerosol size distributions from dental procedures

Aerosol size distributions inherently provide a robust fingerprint of source. Different aerosol sources yield different log-normal distributions with different mean sizes and breadths. Therefore, a detailed analysis and comparison of size distributions from patient procedures and phantom head controls enables identification of potential sources of salivary aerosol. The aerosol size distributions detected from patient procedures are shown alongside phantom head controls on a linear scale (**Supplementary Fig. 2**) and a logarithmic scale (**Supplementary Fig. 3)**. Distributions show that, within each procedure, the patient and phantom head have the same number of modes with similar mode widths (log) and peaks (D_P,C_) but different mode heights (N). Fitted size distributions for ultrasonic scaling, 3-in-1 air + water and slow speed drilling are shown in **Fig. 3**, the remaining procedure fits are presented in **Supplementary Fig. 4**.

**Figure 3.**
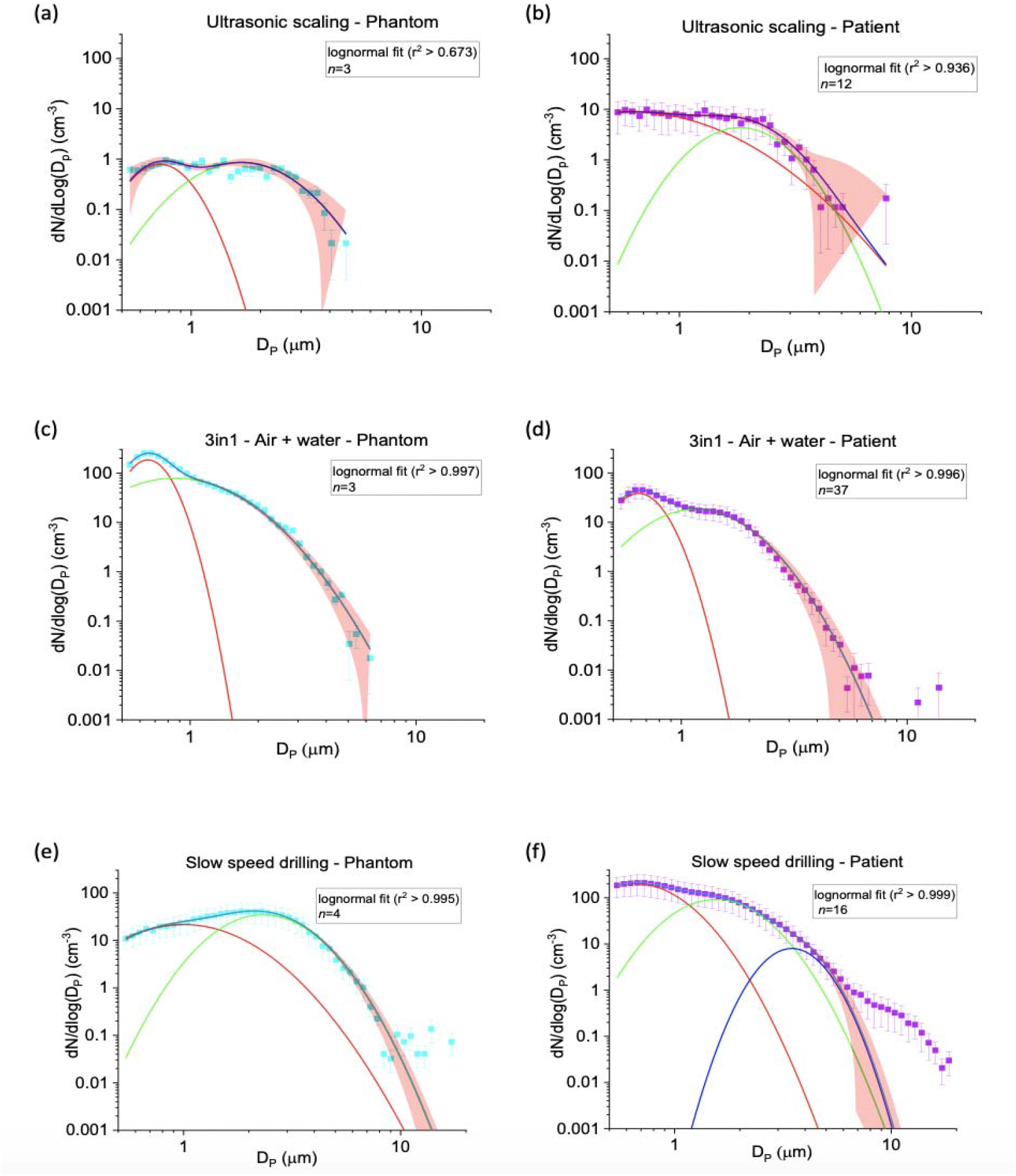
Size distribution data for ultrasonic scaling (a, b), 3-in-1 (c, d) and slow-speed drilling (e, f). Mode 1 (red line), mode 2 (green line), mode 3 (dark blue line) and cumulative bi- or tri-modal fit (blue line). 95% confidence band is shown as the red shaded area.

We assessed how aerosol size distributions from patients may differ from the phantom head control for the same procedure. Such assessment enables identification of sources of salivary aerosol, relative to the non-salivary aerosol generated by the dental instrument. For ultrasonic scaling, bi-modal fits to the patient and phantom head control data show a high level of agreement for the mode width (log(σ)) (Mode 1: *p*= 0.74; Mode 2 *p*= 0.87) and peak (D_P,C_) (Mode 1: *p*= 0.57; Mode 2: *p*= 0.71) between both patient and phantom head control (**Fig. 3a,b** and **Table 2**). This indicates that the phantom head control data (instrument source) may account for all the aerosol seen during ultrasonic patient procedures. Similarly, data from surgical drilling, 3-in-1 air + water and air alone could be represented by bi-modal fits, with shape of size distribution curves similar for both the patient and phantom head controls. Model parameters were similar again, suggesting the aerosol detected arose from the dental instrument source (**Fig. 3c,d, Supplementary Fig. 4a-d, Table 2**).

**Table 2.**
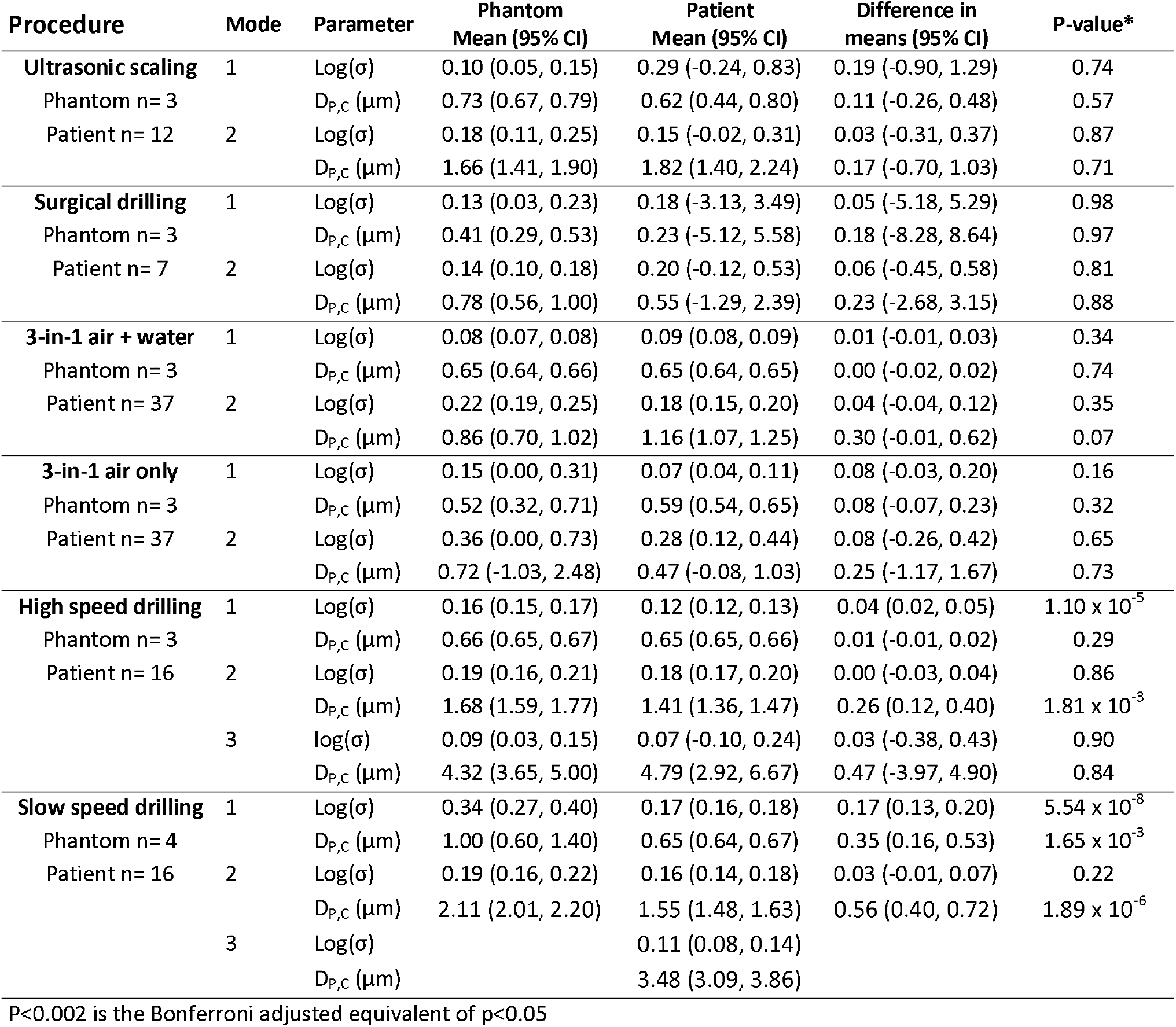
A comparison of aerosol size distribution parameters between the phantom control and patient data.

For high-speed drilling, both the phantom head control and patient data fit can be represented by a tri-modal fit, with similar shaped size distribution curves (**Supplementary Fig. 4e**,**f**). There was statistical evidence passing the multiple testing threshold, that the size distribution modes were different between the phantom head control and the patients (log(σ)) (Mode 1: *p*= 1.10 × 10^−5^) and peak (D_P,C_) (Mode 2: *p* =1.81 × 10^−3^), suggesting the patient aerosol size distribution may not be completely explained by instrument aerosol (**Table 2)**. Slow speed drilling phantom head control data were best represented by a bi-modal fit, whereas three modes were required for the patient data. These aerosol size distributions are different from each other, with clear divergence below 3 µm and above 7 µm particle sizes (**Fig. 3e,f)**. This difference in size distributions was reinforced as the parameters showed strong evidence for a difference in Mode 1 (mean difference log(σ) = 0.17 (95% confidence intervals (95%CI), 0.13, 0.20), *p*= 5.54 × 10^−8^; D_P,C_ = 0.35 (95%CI 0.16, 0.53), *p*= 1.65 × 10^−3^) and Mode 2 (mean difference log(σ) = 0.03 (95%CI -0.01, 0.07), *p*= 0.22; D_P,C_ = 0.56 (95%CI 0.40, 0.72), *p*= 1.89 × 10^−6^) (**Table 2**). Therefore, for slow speed drilling the phantom head control data do not fully explain what was observed during patient procedures.

## Discussion

This study explored aerosol generation during fifteen different dental procedures at source (as close to the patient as possible). Only six procedures generated aerosol detectable above background. Our background particle concentration was very low (0.18 cm^-3^) and of similar magnitude to that generated by a person speaking but less than that generated by a person coughing, enabling confident detection of aerosol produced during dental procedures.

Of the six procedures that generated detectable aerosol, the size distributions observed in patients closely matched those observed in phantom head controls for four of them: ultrasonic scaling, 3-in-1 air/ air + water and surgical drilling. In other words, we did not detect additional aerosol beyond that generated by the dental instrument alone, which is a non-contaminated source. Dental instrument aerosol could in principle be contaminated through impaction and resuspension in the mouth or through coalescence with contaminated aerosol in the oral cavity. However, aerosol coalescence rates within the dental aerosol plume are too small to be significant. For example, coalescence of 1 μm diameter particles at 100 cm^-3^ concentration proceeds with a coagulation coefficient equal to 3.4 × 10^−16^ m^3^ s^-1^, reducing the particle concentration to only 99.999 cm^-3^ in 100 s. Even for coalescence of dissimilar size particles (e.g., 100 nm particles with 1 μm particles), coalescence is so inefficient that the concentration changes by <0.1%. Therefore, if aerosol from the dental instruments cannot pick up patient biological aerosol by coalescence in the oral cavity (either respirable or resulting from the dental procedure), the only remaining alternative is that the aerosol from the dental instrument deposits in the oral cavity and then, having mixed with salivary components and potentially infectious virus, is re-suspended by the instrument. This process would generate an entirely new source of aerosol and be identifiable by the emergence of an additional mode in the size distributions. Because the patient and phantom head size distributions match well for these procedures, it is unlikely the measured aerosol is contaminated by patient biological material unless the new aerosol is generated at a concentration low enough not to be resolved from the size distribution of the instrument-generated aerosol. By contrast, with both high and slow speed drilling there were differences observed between the phantom head and patient aerosol size distributions. The presence of this unexplained aerosol suggests the generation of salivary aerosol and consequently the potential for viral transmission.

Our study in patients supports findings from previous phantom head studies. Din et al. (2021), showed that orthodontic debonding using a high speed drill led to the most significant increase in particles, while combined use of the 3-in-1 air-water syringe did not result in any detectable increase in the aerosol levels. Similarly, Allison et al. (2021) and an N-of-one human volunteer study [22] found that ultrasonic scaling produces mainly instrument-generated aerosol. Similar to these groups, we found the amount of aerosol generated by ultrasonic scaling was low in comparison to high speed drilling (at least 10 times less) and intermittent, with no detectable aerosol for the majority of the time the instrument was in use. This may reflect the non-continuous use of dental instruments, that aerosol does not always escape the oral cavity, that aerosol is mitigated by use of high-volume suction, or that there is directionality to the generated aerosol plume, which cannot be continuously sampled.

Our study characterises aerosol generation during dental procedures but did not test for the presence of SARS-CoV-2, although aerosols and droplets are the vehicles that transmit SARS-CoV-2. Observation of increased aerosol generation does not confirm the potential for pathogen transmission, and it is possible we were unable to identify potential salivary aerosol if the patient size distribution were altered minimally from the phantom control or procedure generated new aerosol at such a low concentration it could not be differentiated from that generated by the dental instrument. While some air sampling studies have detected viable SARS-CoV-2, others have not, and this remains technically challenging [23-25]. The use of time-of-flight mass spectrometry (MALDI-TOF), fluorescein dye or salivary enzyme markers could be useful in determining if unexplained aerosol contains biological material from the patient. For dental instrument-generated aerosol to increase the risk of SARS-CoV-2 transmission, it must interact with saliva containing the virus, be of a size distribution that can contain SARS-CoV-2, withstand irrigant dilution, and ultimately go on to interact with a susceptible host.

In this work, the background aerosol level was low for a typical dental surgery (0.18 cm^-3^), but still 50 times higher than can be achieved in a laminar flow theatre setting (Brown et al. 2021). Very low levels of aerosol (e.g., from breathing or speaking) were not clearly resolved [19, 20]. Nonetheless, we accurately measured aerosol size distributions to identify differences in patient data compared to phantom head controls. Many factors that are uncontrollable in a clinical setting (e.g., patient movement, differences in use of instruments by clinicians, specific tooth or quadrant treated) will affect the aerosol number concentration, but these would minimally affect size distributions. Both sets of experiments are comparable because the set-up for phantom head controls and patient measurements were the same, including relative humidity and temperature.

While we investigated a wide range of dental procedures, it is not clear if these results can be extrapolated to the same instrument being used for a different purpose (e.g., cutting a cavity using a high speed drill) or different instruments performing the same procedure (e.g., piezo surgery instead of surgical drilling). This study limited itself to aerosols in the 0.5–20 μm diameter size range, which includes respirable aerosol. The removal of aerosol in this size range is mainly governed by room ventilation. The studied procedures may generate larger droplets, which tend to behave more ballistically [26]. Particles smaller than those studied here (<0.5 μm diameter) are less likely to harbour the virus [18]. Evaporation may occur between aerosol generation and measurement, potentially altering the aerosol size distribution [27].

It has been suggested that the use of the term AGP should be reconsidered [28]. For instance, coughing can occur during any dental procedure (**Supplementary Figure 5**) and may pose a higher risk of viral transmission than many AGPs because coughing generates orders of magnitude higher aerosol number concentrations than many AGPs [29-31]. The potential for viral transmission may be governed primarily by the proximity of the dental care professional to the patient, given the inevitable exposure to short range aerosol and droplet transmission of respiratory aerosol, rather than by specific dental instruments or procedure. This study provides further evidence for sources of aerosol generation during common dental procedures, enabling a more holistic approach to risk assessment [28].

## Data Availability

Summary level aerosol distribution data can be requested from the authors.

## Author Contributions

T. Dudding, S. Sheikh, F. Gregson, J. Haworth, S. Haworth, A.J. Ireland, J.P. Reid, B.R. Bzdek and M. Gormley contributed to conception and study design. T. Dudding, S. Sheikh, J. Haworth, and M. Gormley were responsible for data acquisition and curation. T. Dudding, S. Sheikh, F. Gregson, and M. Gormley drafted the initial manuscript and all authors contributed to subsequent drafts. Data analysis was carried out by T. Dudding, S. Sheikh, F. Gregson, J. Haworth, S. Haworth, J.P. Reid and B.R. Bzdek. Data interpretation and critical revisions were completed by T. Dudding, S. Sheikh, F. Gregson, J. Haworth, S. Haworth, B.G. Main, A.J. Shrimpton, F.W. Hamilton, A.J. Ireland, N.A. Maskell, B.R. Bzdek, J.P. Reid and M. Gormley and all authors helped write the final version of the manuscript. This study is part of a wider collaboration represented by the AERATOR Group who were responsible for the grant idea, writing and funding of this work. All authors gave final approval and agree to be accountable for all aspects of the work.

## Acknowledgements

Firstly, we would like to thank the patients who volunteered to take part in this study and for recognising the value of this work. We thank all the management, administrative and clinical staff at Bristol Dental Hospital and School for their logistical support with this study. In particular Ms Sarah Bain, Dr Julie Weeks, Ms Sarah Constant, Ms Becki Bullock, Ms Kuldip Bhakerd, Ms Ceit Scott and Mr James Tubman. We also extend our thanks to the nursing staff and clinicians who assisted with the patient procedures, including Dr Alex Gormley, Dr Charlotte Richards and the BSc Dental Hygiene and Therapy students. We would like to thank the Medical Equipment Management Organisation Clinical Engineering (MEMO) team at University Hospitals Bristol and Weston Foundation Trust, Mr John Woods (Nuview Ltd.) for sourcing and supplying the Jade SCA5000C Air Purification units (Surgically Clean Air©, Toronto, Ontario) and Ms Adele Carter, for her assistance with arranging the phantom head simulation equipment. Finally, we acknowledge the AERATOR Group who helped write the grant to obtain the funding for this work.

*AERATOR Group members who contributed to this work include: Alice Milne, James Murray, Johannes Keller, Jules Brown, Andrew Shrimpton, Anthony Pickering, Timothy Cook, Mark Gormley, David Arnold, George Nava, Jonathan Reid, Bryan R Bzdek, Sadiyah Sheikh, Florence Gregson, Fergus Hamilton, Nick Maskell, James Dodd, Ed Moran.

