## Supplement for "A clinical observational analysis of aerosol emissions from dental procedures"

**Supplemental Material**

**Supplementary Methods**

*Environment and equipment*

The Aerodynamic Particle Sizer (APS) (TSI Incorporated, model 3321, Shoreview, NM, USA) measures at a sampling flow rate of 1 L min^-1^ with accompanying sheath flow of 4 L min^-1^. It reports the aerodynamic size of particles in an aerosol plume, size-resolving aerosol number concentration into 52 size bins ranging from 0.5 µm to 20 µm in diameter, with a time integration of 1s. The size bins are equally spaced in log(diameter) space, apart from the smallest size bin (0.5–0.523 µm ). The APS was calibrated by the manufacturer using a standard before use (polystyrene beads of known the size as the calibrant). The APS is regularly returned to the manufacturer (TSI) for calibration and cleaning. Instrument performance over time is monitored by making simultaneous measurements across four different APS instruments in tandem. The reported optical size of the particles was based on an assumed refractive index of pure water at 600 nm wavelength (1.333). All dental treatment was carried out in the same enclosed side room with dimensions measuring 4.27 m x 3.89 m x 2.60 m, with the same set-up for patients and phantom head controls shown in **Supplementary Figure 1**.

Achieving as low background as possible is required to detect potential low level aerosol generating procedures (AGPs) [1]. While we cannot conclude the source(s) of background levels from this study, they may arise from dust, room preparation and cleaning, or human activity before the study began. A clean background environment was created in the treatment room by closing external ventilation vents and using three Jade SCA5000C Air Purification units (Surgically Clean Air©, Toronto, Ontario) on Turbo mode (690 m^3^ h^-1^), with ultrafine particulate filtration using HEPA-Rx. These units were placed in the corners of the room, as far as possible from the dental chair to reduce the background particulate number concentration sufficiently (allowing for sensitive aerosol measurements in the outpatient setting) while not acting to reduce the amount of aerosol being recorded at the sampling funnel.

Owing to the use of the air purifiers, the mean background concentration of 0.18 (+/- SD 0.12) particles cm^-3^ reported by the APS. This was sufficiently low for a baseline cough from each patient to be detected, meaning AGPs could also be identified. UK Infection Prevention Control Guidance standards were followed, including the use of level 3 PPE and FFP3 masks worn by all members of the team, with effective decontamination between patients and a fallow time of 15 mins implemented based on the measured air changes per hour > 10.

Aerosol was sampled into the APS through a funnel and conductive silicon tubing. We conducted an initial pilot study to investigate the optimum position and orientation for the 3D-printed funnel when sampling dental aerosol. This was determined to be 22 cm from soft tissue nasion to the top of the funnel, at approximately 45 degrees on the patient’s left side (11 o’clock position). A 22 cm marked ruler was used to measure between nasion and the top of the funnel at the start of every patient and control procedure (and during the procedure if significant movement occurred), to ensure the correct position. This distance provided adequate space for the clinician to perform the treatment while still being close enough to capture any generated aerosol and was in the path of the exhaled airflow to allow capture of respiratory aerosol.

In the early pilot work, we demonstrated that the transmission efficiency of aerosol sampled through the funnel and subsequent tubing was very high (>99.8% by number, >86% by mass) for aerosol <7 µm diameter [2]. The funnel enables a more robust and sensitive measurement because it intersects a wider cross-section of the dental aerosol plume. Consequently, the minimal transmission losses through the sampling set-up are too small to affect the conclusions of this study. Measurements of particles >7 μm diameter are limited by shot noise (Poisson statistics) owing to their lower relative abundance and therefore quantification in this size range is poorer [1]. We make no correction for these in this study as our primary interest is in quantifying relative rather than absolute risk.

*Patient dental procedure aerosol measurement*

A brief description of the various procedures carried out and equipment used is given below:

- Periodontal treatment: which included examination with a BPE (World Health Organisation) probe, full mouth supragingival scaling or root surface debridement (RSD) of one or two mouth quadrants under local anaesthetic using a Cavitron ® Select ™ SPS ™ (Denstply Sirona, York, USA) ultrasonic scaler (USS) and hand scaling instruments.
- Oral surgery treatment: which included routine dental forceps extractions or the surgical removal of teeth, using a OsseoDoc Drill system (Bien-Air Dental SA, Switzerland) and a straight handpiece (W&H Dentalwerk Bürmoos GmbH, Austria) with round and fissure burs for bone removal, operating at a drive speed of 40,000 rpm with 0.9% saline coolant. Coolant to irrigate the surgical bur was delivered via the peristaltic pump from a 500 mL bag hung on a drip stand.
- Orthodontic debond treatment was performed using band and bracket removers (Ixion ^TM^, DB Orthodontics, Keighley, UK). Both high speed KaVO ^TM^ Bellatorque Mini Lux3 637B (KaVo ^TM^ Dental GmbH, Biberach, Germany) at ≥ 385,000 rpm and slow speed air turbines (KaVo ^TM^ Dental GmbH, Biberach, Germany) at 40,000 rpm, with tungsten carbide debonding burs (8 flute, 0.016 ” diameter, Prima Dental, Gloucester, UK) were used to remove the residual composite or glass ionomer material from each tooth. Each patient had one arch removed with the high speed drill (with water irrigation) and one with the slow drill (no water irrigation) with the arch used for each alternating for each patient. Post-treatment alginate impressions and clinical photographs using cheek retractors and an occlusal mirror were taken.

Wide bore high volume aspiration at 300 L min^-1^ was used during all procedures except oral surgery, for which Medi-Vac^TM^ suction at 60 L min^-1^, with a Yankeur suction tip was used. Detailed protocols were written in Microsoft ® Excel ® with built-in time macros, to accurately time-stamp each procedure (**Appendices 1 – 3**), using a standardised approach. These protocols could easily be adapted as required, given the possibility for *ad hoc* events which may occur as part of the procedure (e.g., patient coughing).

*Phantom head control procedure aerosol measurement*

Where aerosol was detected in patients and where practical, these procedures were repeated in triplicate on a phantom head to mimic the patient treatment as closely as possible. The aerosol size distributions from the phantom head control and patients were compared, with the assumption that if the distributions were the same, all aerosol detected from the patient during the procedure could be explained by the non-salivary contaminated instrument source (represented by the phantom control). Phantom head units were placed in the same position as the patient, using the same equipment and set-up. For the 3-in-1 syringe (air, water, and combined air + water) and removal of residual composite following orthodontic debond with high-speed and slow-speed air turbines, the patient procedure was replicated in the phantom head. For the ultrasonic instrument, it was only practical to replicate supragingival scaling and for the surgical drill, this was used within the phantom head in the same motion and region of the lower third molar teeth as it would be used in a patient, but without contacting the phantom head jaws.

**Statistical analysis**

*Data processing and fitting*

When an instrument was in operation and the APS detected aerosol, the aerosol number concentration and the size distribution data (with fixed size bin of 0.03125 for APS instrument) were extracted for further analysis. The aerosol number concentration and particles per size bin were divided by the sampling number (1 sample was 1 second) to obtain the mean particle number per 1 sample per cm^-3^. For each patient and phantom head control, an aerosol number concentration and particles per size bin were calculated from all the repeats of each instrument signal detected. Therefore, for each phantom head control repeat there would be an n=30 equalling a total of n= 90 for each instrument.

The patient values were combined to generate a procedure aerosol number concentration and were described by medians and interquartile ranges. For each procedure, mean aerosol number concentrations (dN) for a range of particle size bins (D_p_) were calculated by averaging across patients. These were transformed (dN/dLog(D_p_)/cm^-3^) in order to normalise the data (by dividing by the logarithm of the bin width (µm) of 0.03125 (APS)), enabling visual comparison of the size distribution in a standardised form typical for reporting aerosol size distributions. Analysis involved fitting the patient and phantom control size distribution separately for each procedure using **Supplementary Equation 1** to either a bimodal or multimodal fit to determine the best fit by statistical analysis coefficient of determination (r^2^ ) in Origin 2021 (Origin Lab Corporation, Massachusetts). The closer r^2^ was to 1, the better the fit (**Supplementary Figures 2 – 4**).

$$\frac{dN}{dLog\left( D_{P} \right)}=\Sigma\frac{N}{\left( 2\pi\right)^{0.5}\log\left( \sigma\right)}exp\frac{{-(logD_{p}-logD_{P,C})}^{2}}{{2(log\sigma)}^{2}}$$

**Supplementary Equation 1.** The mean APS size distribution data for each instrument was fitted to the multimodal lognormal equation. The D_P_ is the particle diameter (µm), D_P,C_ is the particle diameter at the mode centre, log(σ) relates to the mode width and N is the mode amplitude and related to the sampled concentration (cm^-3^). Fitting was conducted in Origin 2021 (Origin Lab Corporation, Massachusetts).

*Additional time series graphs of treatments*

Example time series for a typical periodontal, oral surgery and orthodontic procedure are included. These are annotated with the time-stamped information as per the protocols (**Appendices 1 – 3**). For example, for ultrasonic scaling shown as A-K, with variability in signal attributed to the fact that the aerosol plume generated was directional and we were sampling at a fixed point (11 o’clock funnel position) (**Supplementary Figures 5 – 7**).

**Supplementary Table 1.** Baseline characteristics of patients for each treatment, by specialty.

|  | Treatment by specialty | | |  |  |
| --- | --- | --- | --- | --- | --- |
|  | Periodontal | Oral surgery | Orthodontic debond | Total | P-value* |
| N (%) | 13 (32) | 11 (27) | 17 (41) | 41 (100) |  |
| Median age (range) | 52 (30 – 75) | 36 (21 – 50) | 18 (18 – 33) | 47 (18 – 75) | < 0.001 |
| Sex (%) |  |  |  |  | 0.852 |
| Male (%) | 6 (46) | 4 (36) | 8 (47) | 18 (44) |  |
| Female (%) | 7 (54) | 7 (64) | 9 (53) | 23 (56) |  |
| Ethnicity |  |  |  |  |  |
| White (%) | 13 (100) | 9 (82) | 15 (88) | 37 (90) |  |
| Asian (%) | 0 (0) | 0 (0) | 1 (6) | 1 (3) |  |
| Black (%) | 0 (0) | 2 (18) | 1 (6) | 3 (7) |  |
| P-value for the test between treatment specialties, one-way ANOVA for age and Fisher’s exact test for sex. Ethnicity has too few values in Asian and Black categories to compare statistically. | | | | | |

**Supplementary Table 2.** Treatment detail for each patient, by specialty.

| Treatment by specialty | Patient no. | Treatment detail |
| --- | --- | --- |
| Periodontal | 1 | Full mouth supragingival scale |
|  | 2 | Full mouth supragingival scale |
|  | 3 | RSD* lower right quadrant |
|  | 4 | RSD lower right quadrant |
|  | 5 | RSD lower right quadrant |
|  | 6 | RSD upper right quadrant |
|  | 7 | RSD upper right and lower right quadrants |
|  | 8 | RSD upper right and lower right quadrants |
|  | 9 | RSD upper right and lower right quadrants |
|  | 10 | RSD upper and lower left quadrants |
|  | 11 | RSD upper left and upper right quadrants |
|  | 12 | RSD upper left quadrant |
|  | 13 | RSD lower left quadrant |
| Oral Surgery | 14 | Surgical removal of teeth UL6 roots, UL8 and LL8 |
|  | 15 | Surgical removal of teeth UL8 and LL8 |
|  | 16 | Extraction of tooth UL6 |
|  | 17 | Surgical removal of teeth UR8 and LR8 |
|  | 18 | Surgical removal of tooth LL8 |
|  | 19 | Surgical removal of teeth UR8 and LR8 |
|  | 20 | Surgical removal of tooth LR8 |
|  | 21 | Surgical removal of tooth LL8 |
|  | 22 | Surgical removal of teeth LL8, UR5 and UR7 |
|  | 23 | Surgical removal of tooth LR8 |
|  | 24 | Extraction of tooth LR6 |
| Orthodontic debond | 25 | Full upper and lower arches |
|  | 26 | Full upper and lower arches |
|  | 27 | Full upper and lower arches |
|  | 28 | Full lower arch |
|  | 29 | Full upper and lower arches |
|  | 30 | Full upper and lower arches |
|  | 31 | Full upper and lower arches |
|  | 32 | Full upper and lower arches |
|  | 33 | Full upper and lower arches |
|  | 34 | Full upper and lower arches |
|  | 35 | Full upper and lower arches |
|  | 36 | Full upper and lower arches |
|  | 37 | Full upper and lower arches |
|  | 38 | Full upper and lower arches |
|  | 39 | Full upper and lower arches |
|  | 40 | Full lower arch |
|  | 41 | Full upper and lower arches |

*RSD (root surface subgingival debridement)

**Supplementary Table 3.** Dental procedures included in the study, the sampling time and percentage time aerosol detected.

| Procedure | Number of measurement repeats | Total sampling time for procedure  (s) | Time aerosol detected above background  (s) | Percentage time aerosol detected above background (%) |
| --- | --- | --- | --- | --- |
| Examination (inc. probing) | 25 | 4020 | 0 | 0 |
| Hand scaling | 3 | 935 | 0 | 0 |
| Routine extraction (non-surgical) | 6 | 1128 | 0 | 0 |
| Soft tissue flap raising | 8 | 1116 | 0 | 0 |
| Bracket removal (debonding and debanding) | 14 | 2098 | 0 | 0 |
| Alginate impressions (upper and lower) | 27 | 1824 | 0 | 0 |
| Local anaesthetic | 37 | 3120 | 0 | 0 |
| Ultrasonic scaling | 12 | 12272 | 1559 | 12.7 |
| 3-in-1 water | 20 | 961 | 0 | 0 |
| 3-in-1 air | 35 | 801 | 199 | 24.8 |
| 3-in-1 air + water | 33 | 772 | 581 | 75.3 |
| High speed drilling | 15 | 3,849 | 1,543 | 40.1 |
| Slow speed drilling | 15 | 3,324 | 1,632 | 49.9 |
| Surgical drilling | 9 | 568 | 316 | 55.6 |
| Suturing | 9 | 1986 | 0 | 0 |

**Supplementary Figure 1.** Photographs of the clinical side room used in the study, with equipment and set-up for phantom head controls.

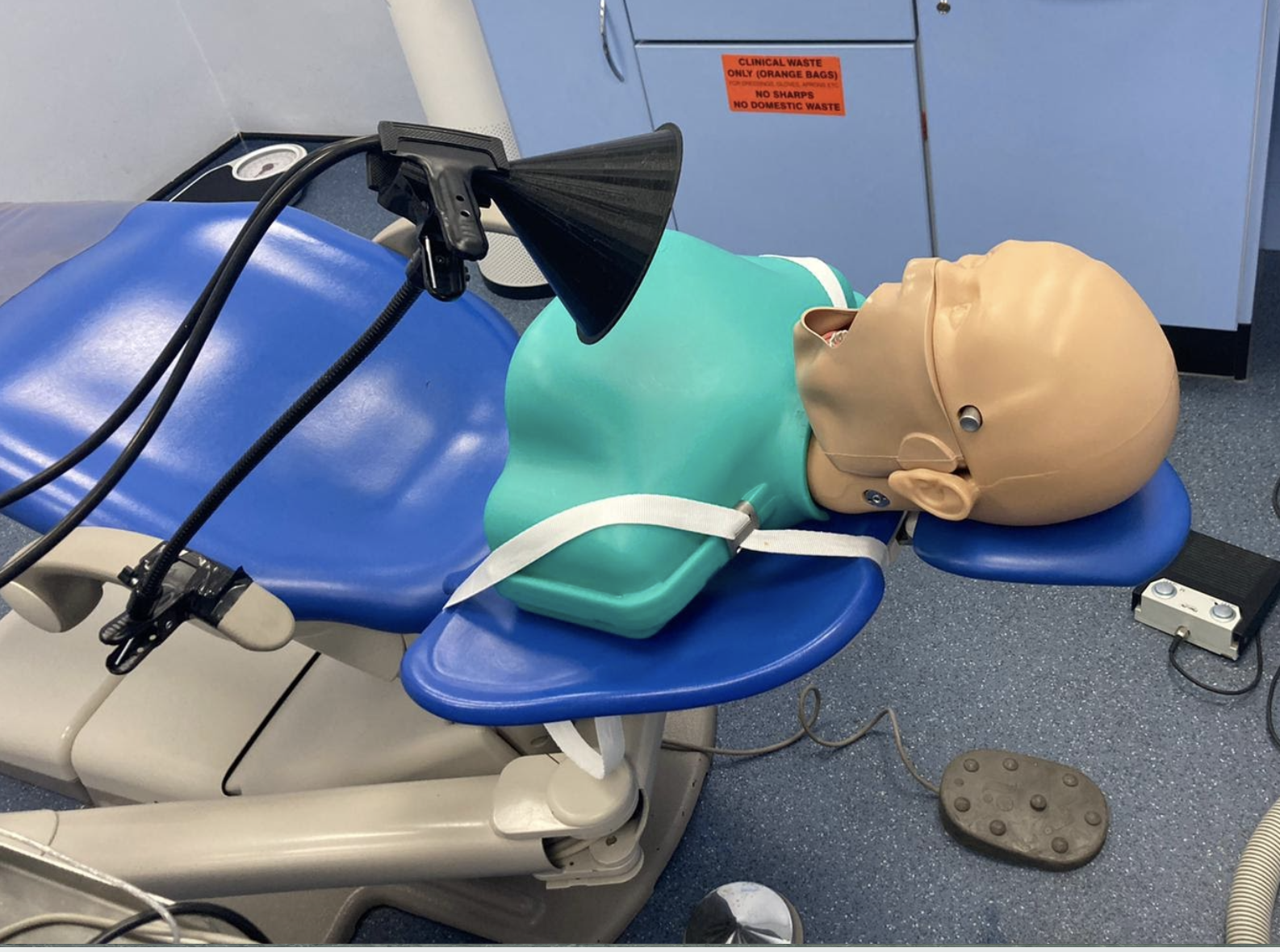

**Supplementary Figure 2.** Aerosol size distributions for dental procedures where aerosol signal was greater than the average background aerosol concentration, plotted with a linear y-axis scale. Both mean patient (purple) and phantom control (light blue) (± standard error) data are presented.

| 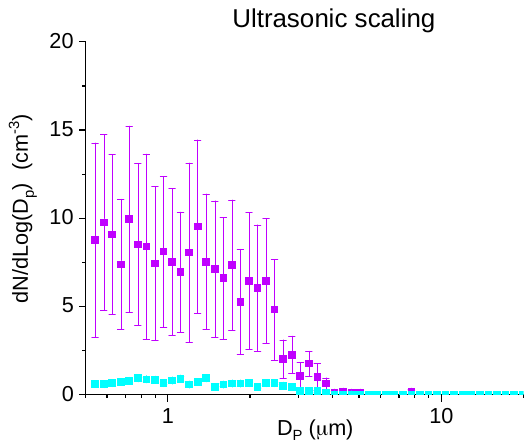  (a) | 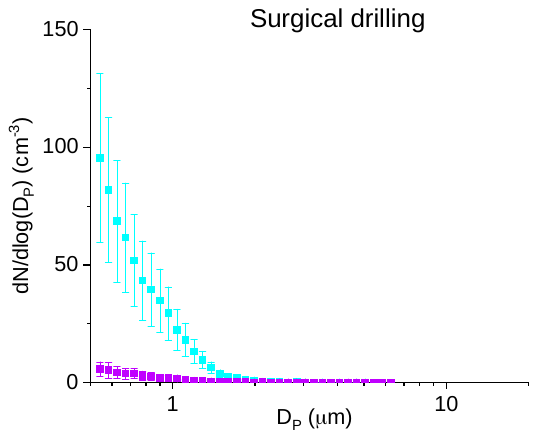  (b) |
| --- | --- |
| 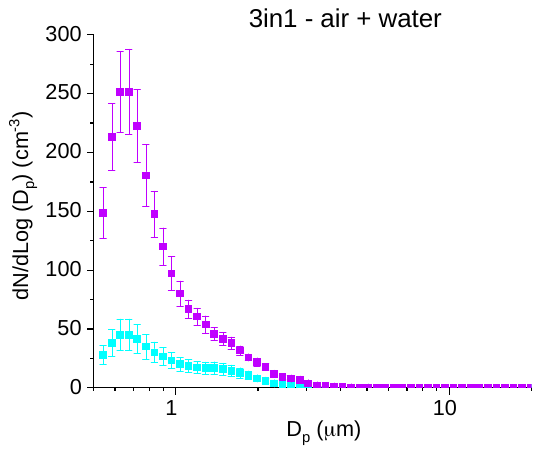  (c) | 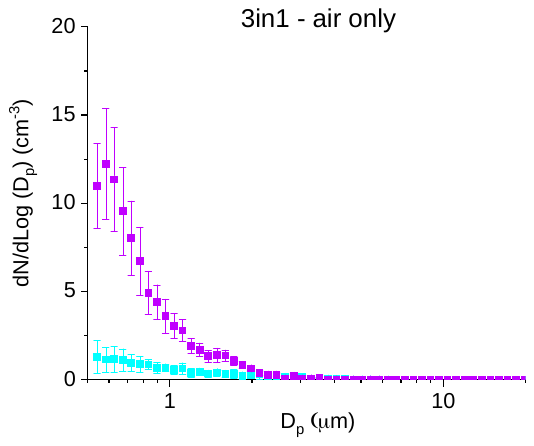  (d) |
| 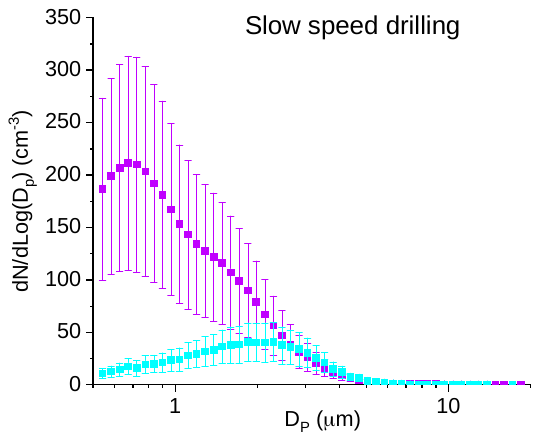  (e) | 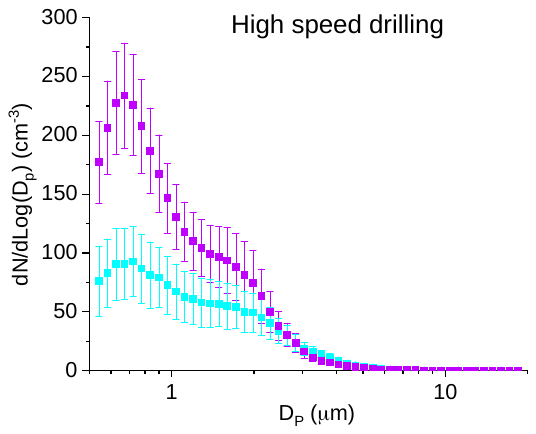  (f) |

**Supplementary Figure 3.** Aerosol size distribution plots with a logarithmic scale for (a-f) dental procedures and (g) patient baseline measurements, where aerosol signal was greater than the average background aerosol number concentration. Both mean patient (purple) and phantom control (light blue) (± standard error) data are presented.

| 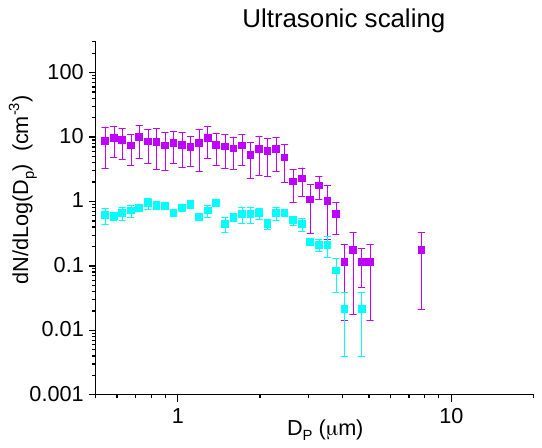  (a) | 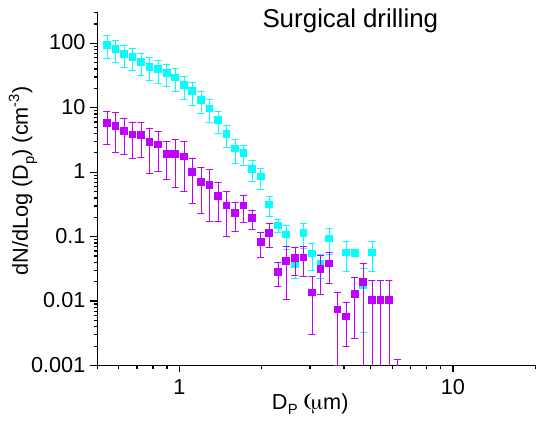  (b) |
| --- | --- |
| 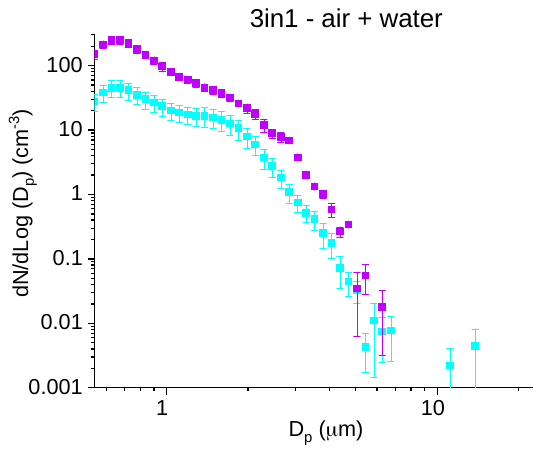  (c) | 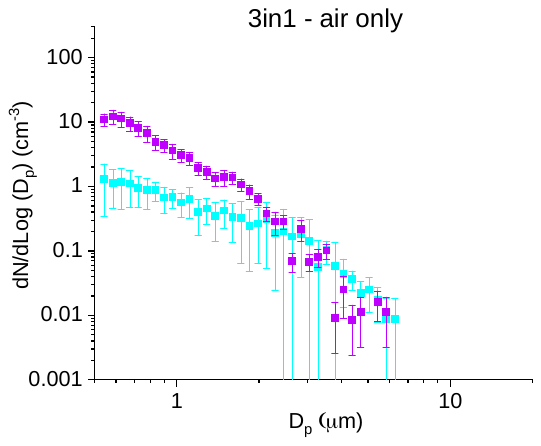  (d) |
| 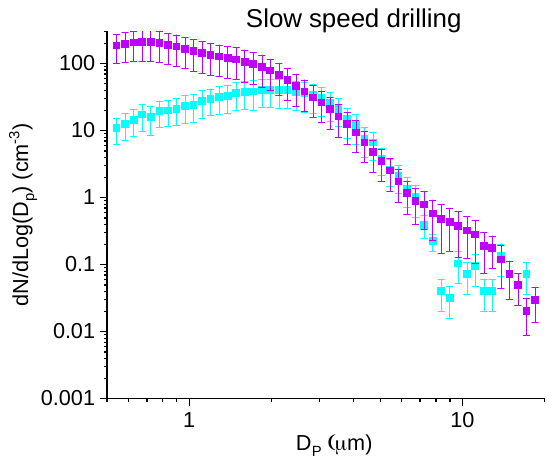  (e) | 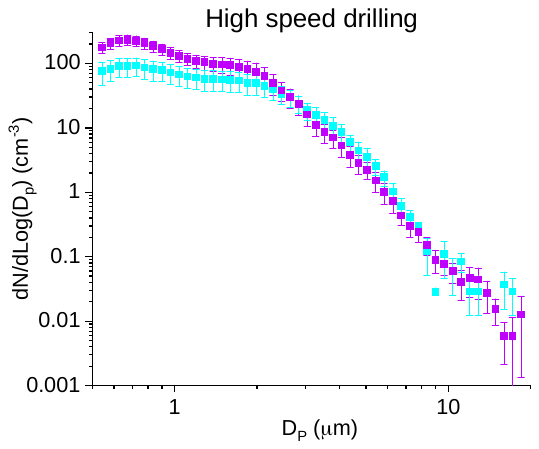  (f) |
| 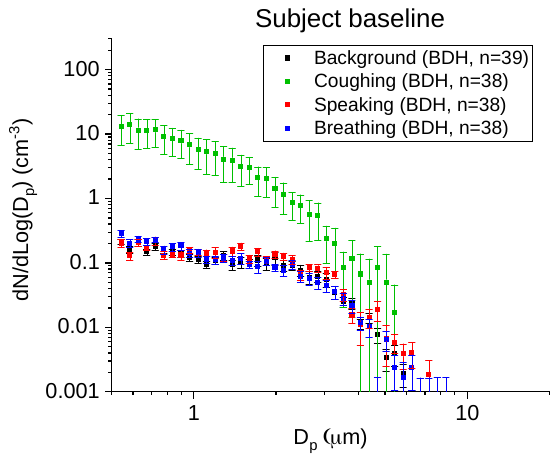  (g) |  |

**Supplementary Figure 4.** Fitting of mean size distribution plots (patient and phantom control) to Supplementary Equation 1. Mode 1 is shown as the red line, mode 2 as the green line, mode 3 (where present) as the dark blue line and the cumulative line of all modes is the mid-blue line. The red area shows the 95% confidence band of the fitting. Fitting correlation r^2^ values are shown.

| 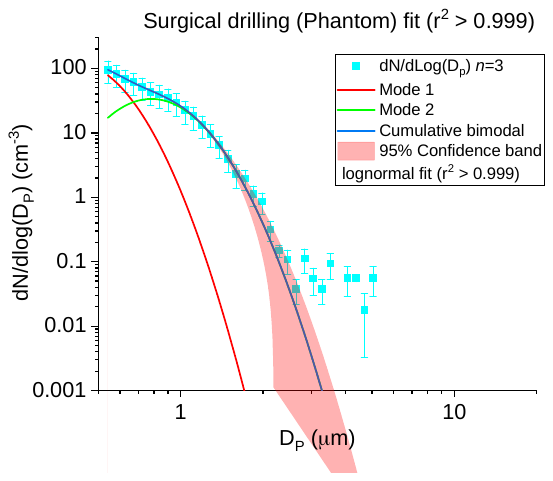  (a) | 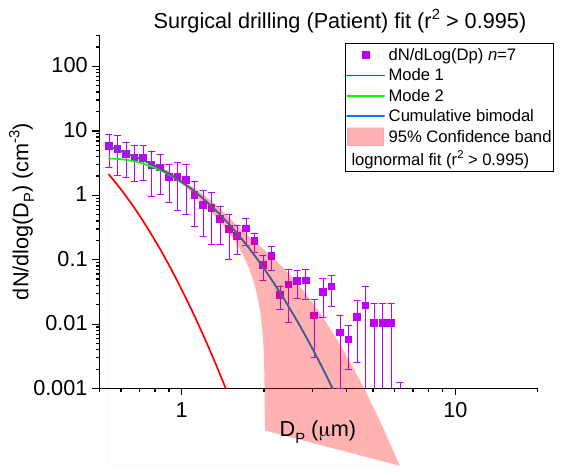  (b) |
| --- | --- |
| 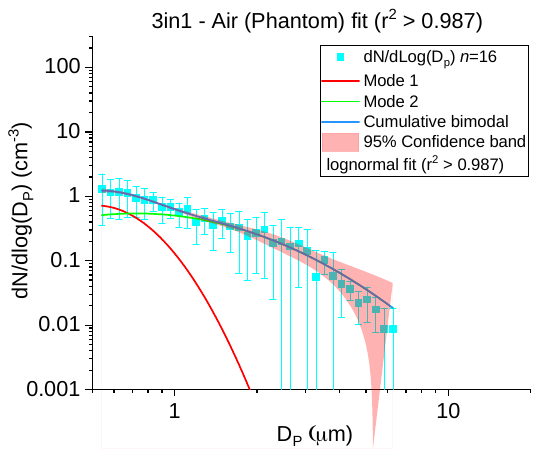  (c) | 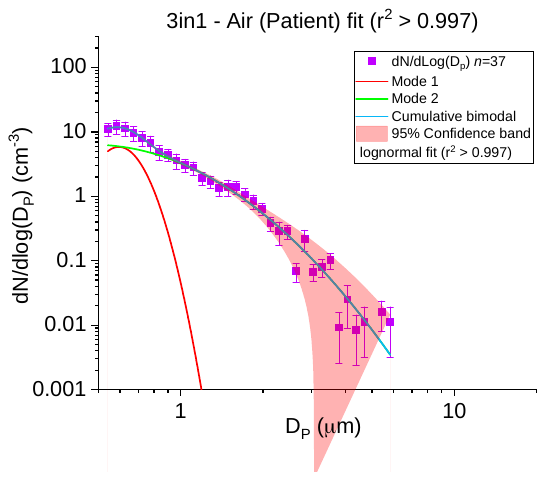  (d) |
| 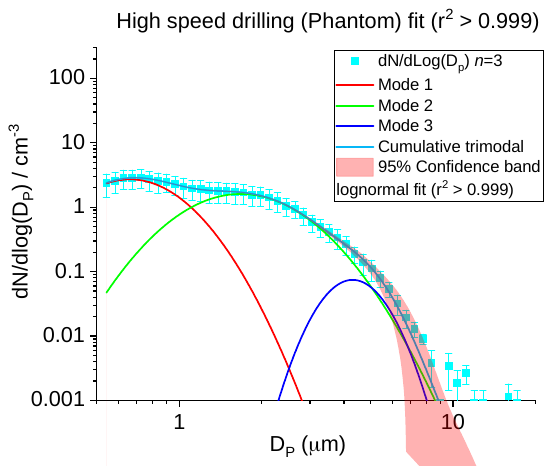  (e) | 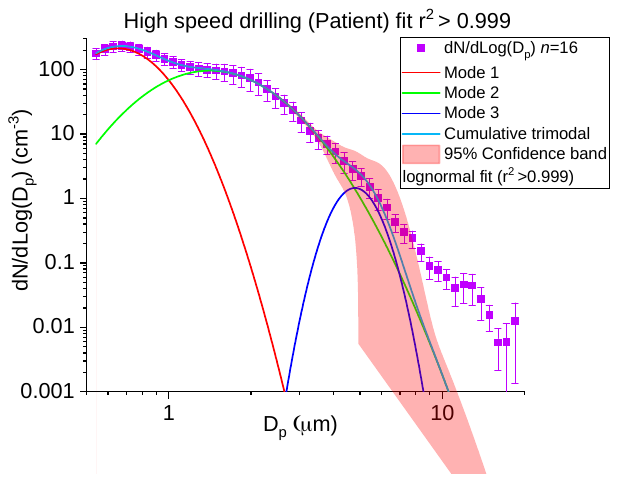  (f) |

**Supplementary Figure 5.** Time series graph for ultrasonic scaling on a patient. Procedures carried out during this treatment were A-K as shown. Area under blue bracket indicates the time period over which the procedure was conducted. The plots show aerosol number concentration over time during the treatment. Inset graph shows baseline measurements (A in main graph). Note that breathing and speaking emit similar or fewer particles than the pre-existing background concentration.

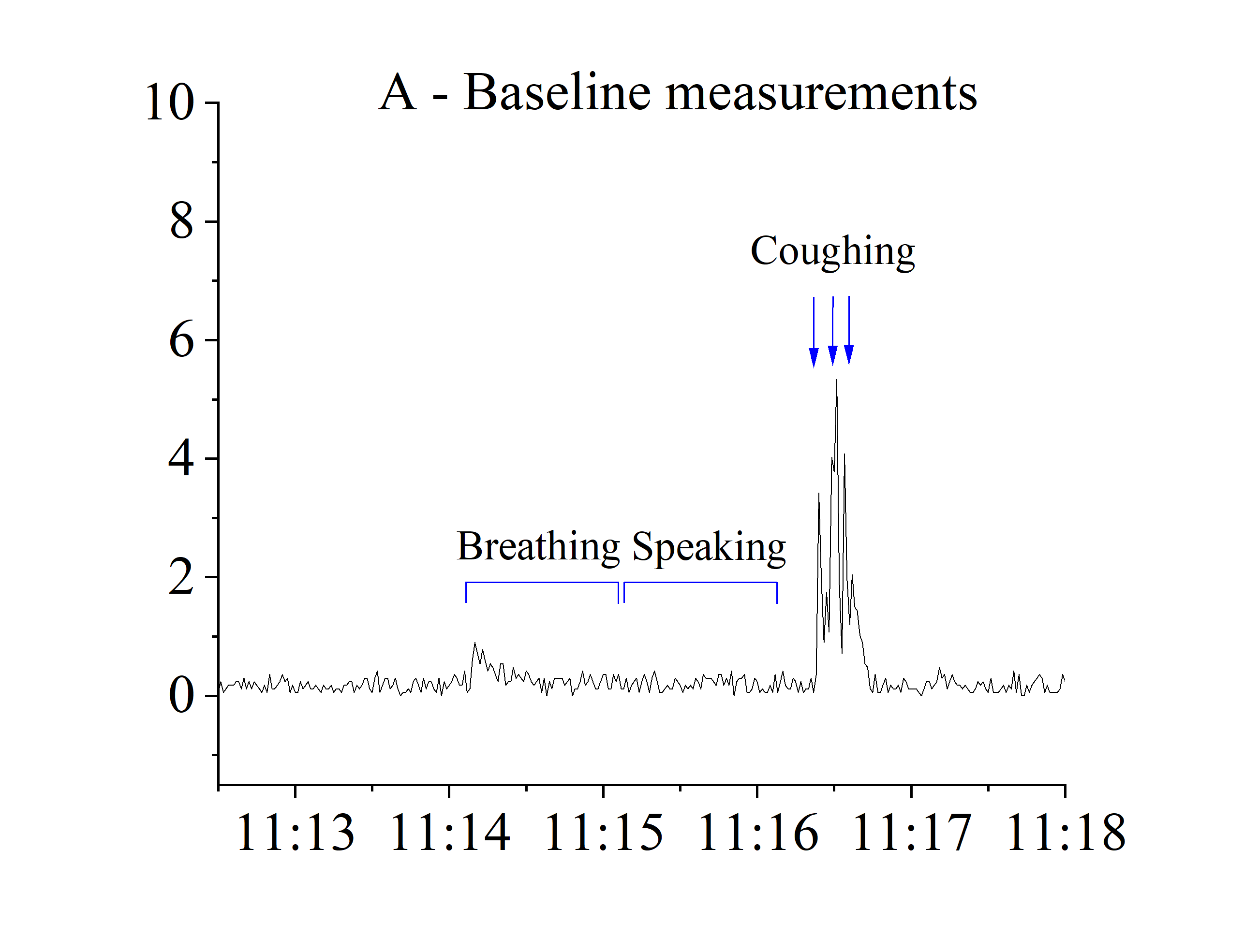

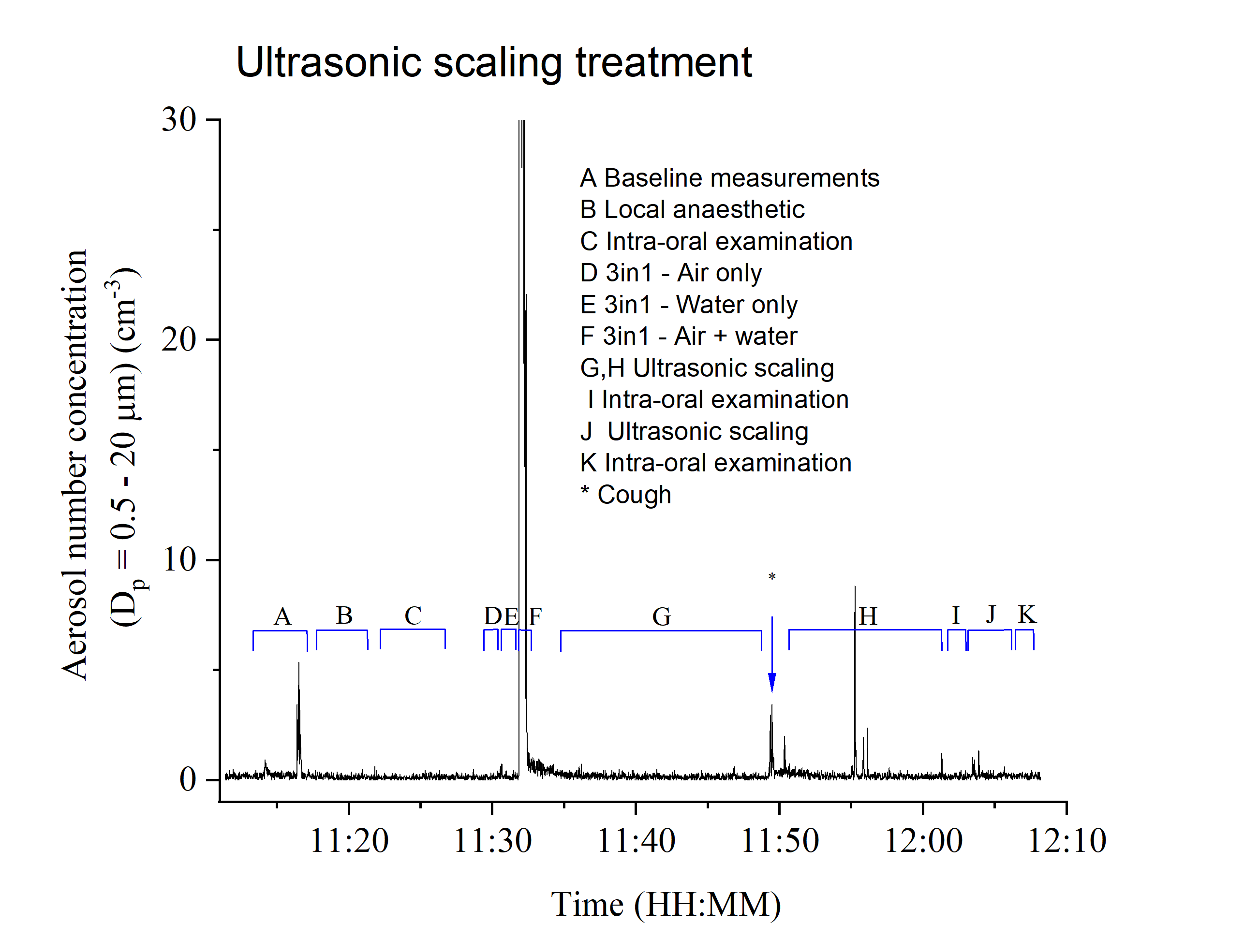

**Supplementary Figure 6.** Time series of surgical removal of a third molar (wisdom) tooth on a patient. Procedures being carried out within a treatment are shown under the blue bracket and described with corresponding alphabetic key.

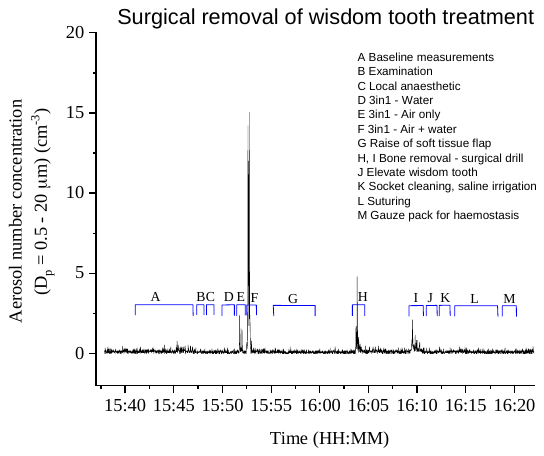

**Supplementary Figure 7.** Time series of an orthodontic debond procedure on a patient. Procedures being carried out within a treatment are shown under the blue bracket and described with corresponding alphabetic key.

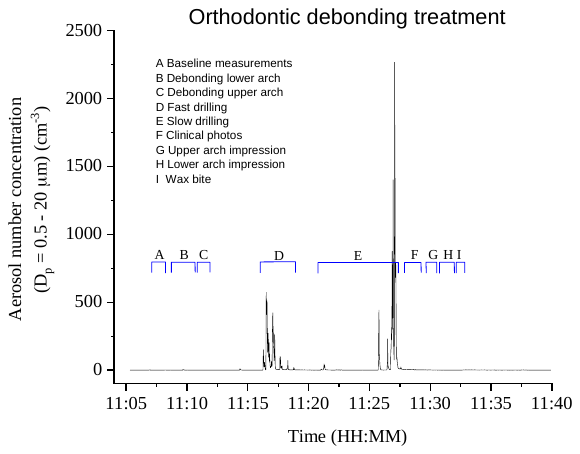

**Appendix 1.** Example of a time-stamped protocol for periodontal treatment

| **Activity** | **Timestamping** | | **Comments** |
| --- | --- | --- | --- |
| Background | Start | 13:41:44 |  |
|  | Stop | 13:45:04 |  |
| Breathing | Start | 13:46:27 |  |
|  | Stop | 13:47:27 |  |
| Speaking | Start | 13:48:00 |  |
|  | Stop | 13:49:00 |  |
| Cough1 | Start | 13:49:31 |  |
| Cough2 | Start | 13:49:55 |  |
| Cough 3 | Start | 13:50:28 |  |
| Examination | Start | 13:51:27 |  |
|  | Stop | 13:52:54 |  |
| 3-in-1 air | Start | 14:00:17 |  |
|  | Stop | 14:00:47 |  |
| 3-in-1 water only (with suction) | Start | 14:01:33 |  |
|  | Stop | 14:02:03 |  |
| 3-in-1 air + water (with suction) | Start | 14:02:35 |  |
|  | Stop | 14:03:05 |  |
| Scale | Start | 14:03:56 | Lower right quadrant |
|  | Stop | 14:08:56 |  |
| Scale | Start | 14:09:27 | Lower left quadrant |
|  | Stop | 14:14:15 |  |
| Scale | Start | 14:14:18 | Lower right quadrant |
|  | Stop | 14:15:45 |  |
| Scale | Start | 14:16:09 | Upper right quadrant |
|  | Stop | 14:20:24 |  |
| Scale | Start | 14:20:27 | Lower anterior teeth |
|  | Stop | 14:22:51 |  |
| Scale | Start | 14:23:09 | Upper left quadrant |
|  | Stop | 14:25:31 |  |
| Intra-oral probing exam | Start | 14:25:48 |  |
|  | Stop | 14:27:27 |  |
| Scale | Start | 14:27:32 | Upper left quadrant |
|  | Stop | 14:28:53 |  |

END OF TREATMENT

**Appendix 2.** Example of a time-stamped protocol for oral surgery treatment

| **Activity** | **Timestamping** | | **Comments** |
| --- | --- | --- | --- |
| Background | Start | 10:31:24 |  |
|  | Stop | 10:35:02 |  |
| Breathing | Start | 10:35:47 |  |
|  | Stop | 10:35:48 |  |
| Speaking | Start | 10:36:00 |  |
|  | Stop | 10:37:00 |  |
| Cough1 | Start | 10:37:32 |  |
| Cough2 | Start | 10:38:02 |  |
| Cough 3 | Start | 10:38:38 |  |
| Examination | Start | 10:52:27 | Check and confirm tooth to be removed |
|  | Stop | 10:54:02 |  |
| Local anaesthetic | Start | 10:54:46 | Left side inferior dental nerve block |
|  | Stop | 10:55:24 |  |
| Local anaesthetic | Start | 10:55:59 | Left side long buccal nerve block plus infiltrations |
|  | Stop | 10:56:32 |  |
| 3-in-1 air | Start | 10:57:00 |  |
|  | Stop | 10:57:30 |  |
| 3-in-1 water only (with suction) | Start | 10:58:03 |  |
|  | Stop | 10:58:33 |  |
| 3-in-1 air + water (with suction) | Start | 10:59:01 |  |
|  | Stop | 10:59:31 |  |
| Raise soft tissue flap | Start | 11:05:18 | Lower left third molar (LL8) tooth |
|  | Stop | 11:05:39 |  |
| Bone removal with surgical drill | Start | 11:06:21 | Around tooth LL8 buccal |
|  | Stop | 11:07:02 |  |
| Bone removal with surgical drill | Start | 11:07:59 | Around tooth LL8 buccal |
|  | Stop | 11:08:49 |  |
| Tooth sectioning using surgical drill | Start | 11:09:39 | Tooth LL8 |
|  | Stop | 11:10:31 |  |
| Elevate sectioned tooth | Start | 11:11:02 |  |
|  | Stop | 11:12:21 |  |
| Socket curettage | Start | 11:14:15 | LL8 socket |
|  | Stop | 11:14:55 |  |
| Saline irrigation | Start | 11:15:21 | LL8 socket |
|  | Stop | 11:16:01 |  |
| Gauze pack placed for haemostasis | Start | 11:16:22 | LL8 socket |
|  | Stop | 11:28:15 | Final examination and check for haemostasis |

END OF TREATMENT

**Appendix 3.** Example of a time-stamped protocol for orthodontic treatment

| **Activity** | **Timestamping** | | **Comments** |
| --- | --- | --- | --- |
| Background | Start | 11:07:37 |  |
|  | Stop | 11:11:33 |  |
| Breathing | Start | 11:11:39 |  |
|  | Stop | 11:12:39 |  |
| Speaking | Start | 11:12:59 |  |
|  | Stop | 11:14:13 |  |
| Cough1 | Start | 11:14:28 |  |
| Cough2 | Start | 11:14:45 |  |
| Cough3 | Start | 11:15:03 |  |
| Examination | Start | 11:15:51 |  |
|  | Stop | 11:17:23 |  |
| Use of debonding pliers and debanding pliers to remove fixed appliances | Start | 11:17:27 | Upper arch |
|  | Stop | 11:19:34 |  |
| Use of debonding pliers and debanding pliers to remove fixed appliances | Start | 11:19:36 | Lower arch |
|  | Stop | 11:21:03 |  |
| 3-in-1 air + water (with suction) | Start | 11:21:19 |  |
|  | Stop | 11:21:49 |  |
| 3-in-1 water only (with suction) | Start | 11:22:12 |  |
|  | Stop | 11:22:42 |  |
| 3-in-1 air | Start | 11:22:57 |  |
|  | Stop | 11:23:27 |  |
| Use of slow speed on upper teeth (to remove composite material) | Start | 11:24:13 | Upper arch composite removal |
|  | Stop | 11:28:43 |  |
| Use of high speed on lower teeth (to remove composite material) | Start | 11:29:03 | Lower arch composite removal |
|  | Stop | 11:31:51 |  |
| Lower arch impression using alginate | Start | 11:36:35 |  |
|  | Stop | 11:37:53 |  |
| Upper arch impression using alginate | Start | 11:38:41 |  |
|  | Stop | 11:39:47 |  |
| Wax Bite recorded | Start | 11:40:09 |  |
|  | Stop | 11:40:24 |  |
| Clinical photographs taken | Start | 11:41:17 |  |
|  | Stop | 11:42:28 |  |

END OF TREATMENT
